# Risk of schizophrenia and bipolar disorder in patients with multiple sclerosis: record-linkage studies

**DOI:** 10.1101/2019.12.09.19014258

**Authors:** Ute-Christiane Meier, Sreeram V Ramagopalan, Michael J Goldacre, Raph Goldacre

## Abstract

**Background:** The epidemiology of psychiatric comorbidity in multiple sclerosis (MS) remains poorly understood.

**Objective:** We aimed to determine the risk of schizophrenia and bipolar disorder in MS patients.

**Material and Methods:** Retrospective cohort analyses were performed using an all-England national linked Hospital Episode Statistics (HES) dataset (1999-2016) and to determine whether schizophrenia or bipolar disorder are more commonly diagnosed subsequently in people with MS (n=128,194), and whether MS is more commonly diagnosed subsequently in people with schizophrenia (n=384,188) or bipolar disorder (n=203,592), than would be expected when compared with a reference cohort (∼15 million people) after adjusting for age and other factors. Adjusted hazard ratios (aHRs) were calculated using Cox proportional hazards models.

**Results:** Findings were dependent on whether the index and subsequent diagnoses were selected as the primary reason for hospital admission or were taken from anywhere on the hospital record. When searching for diagnoses anywhere on the hospital record, there was a significantly elevated risk of subsequent schizophrenia (aHR 1.51, 95% confidence interval (CI) 1.40 to 1.60) and of bipolar disorder (aHR 1.14, 95% CI 1.04 to 1.24) in people with prior-recorded MS and of subsequent MS in people with prior-recorded schizophrenia (aHR 1.26, 1.15-1.37) or bipolar disorder (aHR 1.73, 1.57-1.91), but most of these associations were reduced to null when analyses were confined to diagnoses recorded as the primary reason for admission.

**Conclusion:** Further research is needed to investigate the potential association between MS and schizophrenia and/or bipolar disorder as it may shed light on underlying pathophysiology and help identify potential shared risk factors.

## INTRODUCTION

Multiple sclerosis (MS) is a multifactorial disease of the central nervous system (CNS) characterised by myelin loss, varying degrees of axonal pathology and progressive neurological dysfunction. MS is a classical neuroinflammatory disease, which is caused by immune dysregulation that affects CNS function. Recent insight indicates that similar processes may also play a role in schizophrenia and bipolar disorder and the immune hypothesis in these disorders is receiving growing interest (1).

MS is associated with many neuropsychiatric symptoms, such as depression and anxiety, which even precede MS diagnosis (2) (3). Some reports have suggested a link between MS and schizophrenia and/or bipolar disorder (4).

Several investigators have studied the risk of these conditions in MS patients with no consensus reached. A Canadian study found higher incidence and prevalence estimates of schizophrenia and bipolar disorder in a MS population than in a matched non-MS population (5), another Canadian study reported an association between MS and psychosis (6) and a Danish register-based study found an increased incidence rate ratio of schizophrenia spectrum disorder in MS patients (7). A study on paediatric MS, using the English National Hospital Episode Statistics and mortality data, reported elevated rates of psychotic disorders (RR = 10.76 (2.93-27.63)) (8). However, several studies found no association between schizophrenia and MS. A study from Taiwan found a non-significant increased risk of schizophrenia in patients with MS (9) and another Danish study found no higher than expected prevalence of MS in individuals with schizophrenia (10). Several studies comparing the prevalence of bipolar disorder in MS patients to a comparator population found that bipolar disorder was more common in the MS population (11) (12) (5).

To investigate schizophrenia and bipolar disorder in MS further, we undertook record linkage studies to determine the risk of these disorders in patients with MS using an English National linked Hospital Episode Statistics (HES) dataset.

## METHODS

### Population and data

A Hospital Episode Statistics (HES) dataset covering the population of England from January 1999 to December 2016 was used. The HES data were provided by the NHS Digital (formerly the English national Health and Social Care Information Centre). The Office for National Statistics (ONS) collected data on death registrations in the same time period, which were also supplied to us by NHS Digital. The dataset used in this study, in which successive records for each individual were linked together using personal identifiers irreversibly encrypted by NHS Digital, was constructed by the Oxford record linkage group. (13) Approval for a programme of work covering the construction and analysis of the linked dataset was given by the Central and South Bristol Research Ethics Committee (ref 04/Q2006/176) and has been updated annually.

### Study Design

Retrospective cohort analyses were performed, similar to previous analyses we have undertaken (14) (15). We constructed a cohort of people diagnosed with MS (International Classification of Diseases, Revision 10 (ICD-10) code G35), by identifying the earliest known record of day case care, or inpatient admission, in the dataset for the condition recorded in any diagnostic position in an NHS hospital during the study period. We then searched the database for any subsequent NHS hospital care for, or death from, schizophrenia or bipolar in these cohorts. The ICD codes used to identify schizophrenia or bipolar disorder were, respectively, F20-F29 and F30-31 (10^th^ revision of the ICD). We then repeated the analyses confining the selection criteria to specify that the selected diagnoses were recorded as the main diagnostic reason for the hospital admission. For comparison, a reference cohort was constructed by identifying the earliest known admission for each individual with various other, mainly minor medical and surgical conditions and injuries (appendicectomy, squint, otitis externa, otitis media, haemorrhoids, deflected nasal septum, nasal polyp, impacted tooth and other disorders of teeth, inguinal hernia, in-growing toenail and other diseases of nail, sebaceous cyst, internal derangement of knee, bunion, contraceptive management, cataract, dilation and curettage, selected limb fractures, hip/knee replacement, upper respiratory tract infections, varicose veins). People were included in the MS or reference cohort if they did not have a record of schizophrenia or bipolar either before or at the same time as the first record of MS or the reference condition. We also looked to see if associations were present in the other direction chronologically i.e. whether hospitalisation for MS was increased after hospitalisation for schizophrenia or bipolar disorder.

### Statistical methods

Taking the example of schizophrenia after MS, we used Cox proportional hazards methods to calculate hazard ratios in order to compare the incidence of schizophrenia in the MS cohort with the reference cohort after adjustment for age (in five-year age groups), sex, calendar year of first recorded admission, region of residence, quintile of patients’ Index of Deprivation score (as a measure of socio-economic status), and total number of hospital admissions (excluding those that contained the outcome diagnosis i.e. in this example schizophrenia). Entry to the cohort was the index date of first known hospital admission for MS (or any one of the reference conditions). Exit was earliest known date of hospital-recorded schizophrenia, or date of death, or 31 Dec 2017 (whichever occurred first).

We further subdivided the outcomes into people whose earliest known schizophrenia record was less than one year after the earliest known record of MS, and people whose earliest known schizophrenia record was more than one year after the earliest known record of MS. Our reasoning in doing so was to reduce the possibility of including some short-term associations that might have resulted from misdiagnosis of one condition for the other.

## RESULTS

There were 128,194 people in the MS cohort (65,510 with the condition recorded as primary diagnosis), 384,188 people in the schizophrenia cohort (227,807 as primary diagnosis), and 203,592 people in the bipolar disorder cohort (94,372 as primary diagnosis). There were 15,049,357 people in the reference cohort.

For each analysis, observed numbers of people with each disease outcome and adjusted hazard ratios are reported in the Table.

### Risk of schizophrenia (primary or subsidiary diagnosis)

There was a significantly elevated hazard ratio for schizophrenia after hospital admission for MS (aHR= 1.51, 95% CI 1.40-1.62), both within a year (aHR= 1.75, 95% CI 1.46 to 2.10) and more than a year after hospital admission for MS (aHR= 1.47, 95% CI 1.35 to 1.59). The hazard ratio for MS after hospital admission for schizophrenia was also significantly increased but, overall, lower in magnitude than that seen for schizophrenia after MS (aHR= 1.26, 95% CI 1.15 to 1.37).

### Risk of bipolar disorder (primary or subsidiary diagnosis)

There was a modest but significantly increased hazard ratio for bipolar disorder after hospital admission for MS (aHR= 1.14, 95% CI 1.04 to 1.24), which reduced when looking more than a year after hospital admission for MS (aHR= 1.06, 95% CI 0.96 to 1.16). The hazard ratio for MS after hospital admission for bipolar disorder was significantly increased (aHR= 1.73, 95% CI 1.57 to 1.91) and persisted when excluding patients first admitted with MS within 1 year of first admission with MS (aHR= 1.65, 95% CI 1.48 to 1.84).

### Restriction of analyses to diagnoses recorded as the main reason for admission

This restriction attenuated the estimates of risk towards null. Most dropped below the 5% level of statistical significance. The association between prior bipolar disorder and subsequent multiple sclerosis remained significant, at a time interval of at least a year after first bipolar admission, with HR= 1.29 (95% CI 1.05 to 1.60).

## DISCUSSION

### Main findings

We analysed records from an English national dataset of linked Hospital Statistics and mortality statistics and found that people admitted to hospital with MS coded anywhere on the hospital record had a significantly increased subsequent rate of occurrence of schizophrenia and bipolar disorder coded anywhere on the record. When we reversed the chronological sequence of the diseases, in the equivalent analysis of diagnoses coded anywhere on the record, we found that the risk of MS after a hospital admission for schizophrenia and bipolar disorder was also significantly increased. Most of the associations reported were no longer significant when the diagnoses were restricted to the main reasons for hospital admission, i.e. omitting diagnoses recorded as incidental.

### Comparisons with other studies

Taking the diagnoses from anywhere on the hospital record, the observed increased risk of schizophrenia in MS patients is in line with a Danish study, which found an increased risk (RR=1.44, 95% CI 1.03-1.94) (7). A recent Taiwanese study reported a hazard ratio (HR) of 2.21 (95% CI 0.52-9.34) for schizophrenia after MS, similar to our rate ratio of 2.08, but the Taiwan finding was not statistically significant (9).

Taking the diagnoses from anywhere on the hospital record, we also found an increased risk of MS after a hospital admission for schizophrenia. A previous Danish study also reported an elevated incidence rate for MS in patients with schizophrenia spectrum disorder (RR= 1.57, 95% CI 1.29-1.90); however, in this study the incidence stayed increased five or more years after onset of schizophrenia (RR= 1.53, 95% CI 1.18-1.95) (16). On the contrary, a Swedish study found an inverse association with a decreased risk of MS in patients with schizophrenia (HR= 0.6, 95% CI 0.4-0.9) (17).

Again taking the diagnoses from anywhere on the hospital record, our study also found an increased risk of bipolar disorder in patients with MS, and an increased risk of MS in patients with bipolar disorder, findings which are in line with a Canadian study which observed an increased risk of incident bipolar disorder in patients with MS (HR= 2.67, 95% CI 2.29-3.11) (11), and a Swedish study which observed an increased risk of MS in patients with bipolar disorder (HR= 1.8, 95% CI 1.6-2.2) (17). In our study, the increased risk of MS in patients with bipolar disorder remained when the analyses were restricted to diagnoses recorded as the main reason for the hospital admissions.

### Interpretation

The associations in our study between MS, schizophrenia and bipolar disorder were sensitive to whether or not the diseases were searched for as the primary reason for hospital admission, and as such may only cautiously be interpreted as evidence of disease mechanisms shared by these conditions. MS is a classical neuroinflammatory disease, however, recent evidence highlights that similar processes such as microglial activation, pro-inflammatory cytokines, molecular mimicry between pathogens and brain antigens, anti-neuronal antibodies, self-reactive T-cells and disturbances of the blood-brain barrier are involved in psychiatric diseases such as schizophrenia and bipolar disorder (1). Other possible explanations include shared unmeasured confounders and the possibility of surveillance bias. On confounders, we took account of age, sex, deprivation, and other factors described in Methods; but the possibility exists of other confounders not available in the dataset. Furthermore, surveillance bias could be a factor: if patients with, say, MS are under continuing medical care, this may lead to a higher probability than that in controls (i.e. in the reference cohort) that they might be diagnosed and hospitalised with, say, schizophrenia. The fact that the associations were most evident when incidental diagnoses were included supports the possibility of a surveillance effect.

### Strength and limitations

The strength of the study includes the use of a large, national population based dataset. However, we were wholly dependent on the reliability of the coded diagnostic data in the dataset. The associations may also be overestimated because any bias of association may be driven by severe presentation as it is based on hospital records. Treatment may also complicate the findings and was not taken into account. For example, we are aware that steroid treatment, which is often used to treat a MS relapse, can induce psychosis. In all, our findings should therefore be regarded as preliminary and hypothesis generating rather than definitive.

### Conclusions

People admitted to hospital with MS may be at an increased risk of schizophrenia and bipolar disorder and patients admitted to hospital with bipolar disorder and schizophrenia may be at an increased risk of MS. Clinicians treating patients with these conditions should be aware of these possible co-occurring morbidities. However, there are some inconsistencies in our findings which we describe but cannot fully explain. The findings should be regarded as indicating possible associations and as speculative rather than definitive but, if confirmed, they may indicate a role for a common aetiology underlying these neuropsychiatric disorders.

## Data Availability

The data is available upon request

---

**Table:** Associations between multiple sclerosis (MS) and schizophrenia (SZ) or bipolar disorder (BP), English national Hospital Episode Statistics, 1999-2016

| Exposure | Outcome | Follow-up interval | Exposure and outcome taken from anywhere on the record |  |  |  |  | Exposure and outcome as primary diagnosis only |  |  |  |  |
| --- | --- | --- | --- | --- | --- | --- | --- | --- | --- | --- | --- | --- |
|  |  |  | Total | Observed | HR | 95%CI | p value | Total | Observed | HR | 95%CI | p value |
| BP | MS | All | 203592 | 399 | 1.73 | (1.57-1.91) | 0.0000 | 94372 | 103 | 1.34 | (1.11-1.63) | 0.0029 |
| MS | BP |  | 128194 | 531 | 1.14 | (1.04-1.24) | 0.0035 | 65510 | 77 | 0.86 | (0.69-1.08) | 0.1960 |
| SZ | MS |  | 384188 | 499 | 1.26 | (1.15-1.37) | 0.0000 | 227807 | 130 | 0.82 | (0.69-0.98) | 0.0299 |
| MS | SZ |  | 128194 | 719 | 1.51 | (1.4-1.62) | 0.0000 | 65510 | 142 | 1.11 | (0.94-1.31) | 0.2010 |
| BP | MS | <1yr | 203592 | 69 | 2.36 | (1.85-3.01) | 0.0000 | 94372 | 14 | 1.79 | (1.05-3.05) | 0.0315 |
| MS | BP |  | 128194 | 91 | 1.75 | (1.42-2.16) | 0.0000 | 65510 | 16 | 1.32 | (0.8-2.16) | 0.2736 |
| SZ | MS |  | 384188 | 95 | 2.02 | (1.64-2.49) | 0.0000 | 227807 | 19 | 1.22 | (0.77-1.93) | 0.3997 |
| MS | SZ |  | 128194 | 117 | 1.75 | (1.46-2.1) | 0.0000 | 65510 | 33 | 1.79 | (1.26-2.52) | 0.0010 |
| BP | MS | 1yr+ | 177633 | 330 | 1.65 | (1.48-1.84) | 0.0000 | 88576 | 89 | 1.29 | (1.05-1.6) | 0.0160 |
| MS | BP |  | 114150 | 440 | 1.06 | (0.96-1.16) | 0.2333 | 60155 | 61 | 0.79 | (0.61-1.02) | 0.0676 |
| SZ | MS |  | 335972 | 404 | 1.15 | (1.04-1.27) | 0.0055 | 212920 | 111 | 0.78 | (0.65-0.94) | 0.0099 |
| MS | SZ |  | 114135 | 602 | 1.47 | (1.35-1.59) | 0.0000 | 60140 | 109 | 1.00 | (0.83-1.21) | 0.9985 |

## REFERENCES

1. Pape K, Tamouza R, Leboyer M, Zipp F. Immunoneuropsychiatry - novel perspectives on brain disorders. Nat Rev Neurol. 2019;15(6):317-28.

2. Disanto G, Zecca C, MacLachlan S, Sacco R, Handunnetthi L, Meier UC, et al. Prodromal symptoms of multiple sclerosis in primary care. Ann Neurol. 2018;83(6):1162-73.

3. Marrie RA, Reingold S, Cohen J, Stuve O, Trojano M, Sorensen PS, et al. The incidence and prevalence of psychiatric disorders in multiple sclerosis: a systematic review. Mult Scler. 2015;21(3):305-17.

4. Jeppesen R, Benros ME. Autoimmune Diseases and Psychotic Disorders. Front Psychiatry. 2019;10:131.

5. Marrie RA, Fisk JD, Tremlett H, Wolfson C, Warren S, Tennakoon A, et al. Differences in the burden of psychiatric comorbidity in MS vs the general population. Neurology. 2015;85(22):1972-9.

6. Patten SB, Svenson LW, Metz LM. Psychotic disorders in MS: population-based evidence of an association. Neurology. 2005;65(7):1123-5.

7. Benros ME, Nielsen PR, Nordentoft M, Eaton WW, Dalton SO, Mortensen PB. Autoimmune diseases and severe infections as risk factors for schizophrenia: a 30-year population-based register study. Am J Psychiatry. 2011;168(12):1303-10.

8. Pakpoor J, Goldacre R, Schmierer K, Giovannoni G, Waubant E, Goldacre MJ. Psychiatric disorders in children with demyelinating diseases of the central nervous system. Mult Scler. 2018;24(9):1243-50.

9. Wang LY, Chen SF, Chiang JH, Hsu CY, Shen YC. Autoimmune diseases are associated with an increased risk of schizophrenia: A nationwide population-based cohort study. Schizophr Res. 2018;202:297-302.

10. Eaton WW, Byrne M, Ewald H, Mors O, Chen CY, Agerbo E, et al. Association of schizophrenia and autoimmune diseases: linkage of Danish national registers. Am J Psychiatry. 2006;163(3):521-8.

11. Marrie RA, Patten SB, Greenfield J, Svenson LW, Jette N, Tremlett H, et al. Physical comorbidities increase the risk of psychiatric comorbidity in multiple sclerosis. Brain Behav. 2016;6(9):e00493.

12. Jun-O’Connell AH, Butala A, Morales IB, Henninger N, Deligiannidis KM, Byatt N, et al. The Prevalence of Bipolar Disorders and Association With Quality of Life in a Cohort of Patients With Multiple Sclerosis. J Neuropsychiatry Clin Neurosci. 2017;29(1):45-51.

13. Goldacre M, Kurina L, Yeates D, Seagroatt V, Gill L. Use of large medical databases to study associations between diseases. QJM. 2000;93(10):669–75.

14. Goldacre MJ, Duncan ME, Griffith M, Cook-Mozaffari P. Psychiatric disorders certified on death certificates in an English population. Soc Psychiatry Psychiatr Epidemiol. 2006;41(5):409–14.

15. Seminog OO, Goldacre MJ. Risk of pneumonia and pneumococcal disease in people hospitalized with diabetes mellitus: English record-linkage studies. Diabet Med. 2013;30(12):1412–9.

16. Benros ME, Pedersen MG, Rasmussen H, Eaton WW, Nordentoft M, Mortensen PB. A nationwide study on the risk of autoimmune diseases in individuals with a personal or a family history of schizophrenia and related psychosis. Am J Psychiatry. 2014;171(2):218–26.

17. Johansson V, Lundholm C, Hillert J, Masterman T, Lichtenstein P, Landen M, et al. Multiple sclerosis and psychiatric disorders: comorbidity and sibling risk in a nationwide Swedish cohort. Mult Scler. 2014;20(14):1881–91.

